# Loss of Y in leukocytes as a risk factor for critical COVID-19 in men

**DOI:** 10.1101/2022.01.19.22269521

**Authors:** Bożena Bruhn-Olszewska, Hanna Davies, Daniil Sarkisyan, Ulana Juhas, Edyta Rychlicka-Buniowska, Magdalena Wójcik, Monika Horbacz, Marcin Jąkalski, Paweł Olszewski, Jakub O. Westholm, Agata Smialowska, Karol Wierzba, Åsa Torinsson Naluai, Niklas Jern, Lars-Magnus Andersson, Josef D. Järhult, Natalia Filipowicz, Eva Tiensuu Janson, Sten Rubertsson, Miklós Lipcsey, Magnus Gisslén, Michael Hultström, Robert Frithiof, Jan P. Dumanski

## Abstract

COVID-19 shows an unexplained, strong male bias for severity and mortality. Loss of Y (LOY) in myeloid cells is a risk factor candidate in COVID-19 because of associations with many age-related diseases and its effect on transcription of immune genes. We report the highest levels of LOY in cells that are crucial for the development of severe COVID-19 phenotype, such as low-density neutrophils, granulocytes, and monocytes reaching 46%, 32%, and 29%, respectively, from men with critical COVID-19 (n=139). LOY in sorted subpopulations of leukocytes correlated with increased thrombocyte count, thromboembolic events, invasive mechanical ventilation and a history of vessel disease. In recovered patients, LOY decreased in whole blood and peripheral blood mononuclear cells. Moreover, sc-RNA-seq analysis of CD14+ monocytes from 30 COVID-19 males and 34 controls revealed pervasive transcriptional downregulation in LOY-cells, notably affecting HLA class I and II genes important for antigen presentation. The data support a link between LOY and emergency myelopoiesis as well as the role of LOY in modulation of COVID-19 severity. Our results might also be relevant for other viral infections showing similar male bias.

---

Leukocytes from aging males frequently show mosaic loss of chromosome Y (LOY) [1, 2]. It is detectable in whole blood DNA from >40% of the men above the age of 70 years [3] and reaches 57% in the analysis of 93-year-old men, indicating an increasing frequency with age [4]. Recent single-cell analyses of peripheral blood mononuclear cells (PBMCs) from 29 aging men (median age 80 years, range 64-94 years) identified cells with LOY in every studied subject, making this the most common post-zygotic mutation [5]. In serial analysis of male blood samples LOY is a dynamic process [6]. LOY is most common in leukocytes, but has also been shown in other tissues although with considerably lower frequencies [4, 7]. Risk factors for LOY include age, smoking and germline predisposition [1-3, 8]. LOY in whole blood has been associated with increased risk for all-cause mortality, hematological and non-hematological cancers and other age-related disorders such as Alzheimer’s disease, diabetes and cardiovascular events, among others [1, 2, 7, 9-14]. Thus, carriers of LOY have an increased risk for diseases both inside and outside of the hematopoietic system and the mechanism(s) behind these associations largely remain to be elucidated. Recent studies suggest that LOY could have a direct physiological role through **<u>L</u>**OY-**<u>a</u>**ssociated **t**ranscriptional **<u>e</u>**ffects (LATE) on global gene expression in a pleiotropic manner as well as being a biomarker of genomic instability in somatic tissues [5]. Moreover, dysregulation of immune genes, including pathways related to viral life-cycle, was pronounced in LOY-cells [5, 15].

Coronavirus disease 2019 (COVID-19) is caused by the new SARS-CoV-2 virus [16]. Critically affected patients present with bilateral pneumonia, in many cases progressing to acute respiratory distress syndrome with a strong decrease in pulmonary function requiring treatment with non-invasive or invasive mechanical ventilation [17]. Results suggest that neutrophil activation is a hallmark of COVID-19 together with lymphocytopenia and that patients with the critical disease show substantial elevation of circulating neutrophils [18-20]. In COVID-19 patients, neutrophils adopt a so-called low-density phenotype, when they are activated and prone to spontaneous release of neutrophil extracellular traps (NETs) in capillaries. Intra-vascular aggregations of NETs lead to occlusion of the affected vessels, disturbed microcirculation and organ damage. Furthermore, hyper-coagulation and thromboembolic events are also contributing to the mortality from COVID-19 [21, 22]. These symptoms are possibly a consequence of dysfunction in myeloid cell lineages and growing evidence corroborate this hypothesis [23, 24]. Another feature of the disease is a strong and unexplained bias of males who are critically ill and die from COVID-19. Of 7874 patients treated and registered since March 2020 at Intensive Care Units (ICU) in Sweden, 70.1% were men (Swedish Intensive Care Database [SIR], data from September 9, 2021). Other studies of COVID-19 patients confirm this strong male bias [25, 26] and this aspect of COVID-19 pathogenesis is understudied [27]. Thus, there might exist a so far unidentified male-specific risk factor, predisposing men for a life-threatening disease course of COVID-19. We hypothesized that LOY in cells from myeloid lineage might be linked to the development of the critical disorder and predominantly studied here critically ill COVID-19 males.

## Myeloid lineage cells from COVID-19 patients show the highest levels of LOY

We initially reanalyzed the publicly available single-cell RNA sequencing dataset [18], adding LOY-status for all sequenced cells as described [5] (**Fig. 1** and **Fig. S1**). All critically ill COVID-19 patients (WHO grade >=5) described in Schulte-Schrepping et al, 2020 were included; i.e. 6 male patients with a total of 9 samples (three patients were sampled twice) taken during ICU stay. In this re-analysis, we used two previously published cell type annotations [18, 28] with concordant results. Neutrophils, immature neutrophils (also called low-density neutrophils, LDNs) and other myeloid cells clearly showed the highest levels of LOY.

**Fig. 1.**
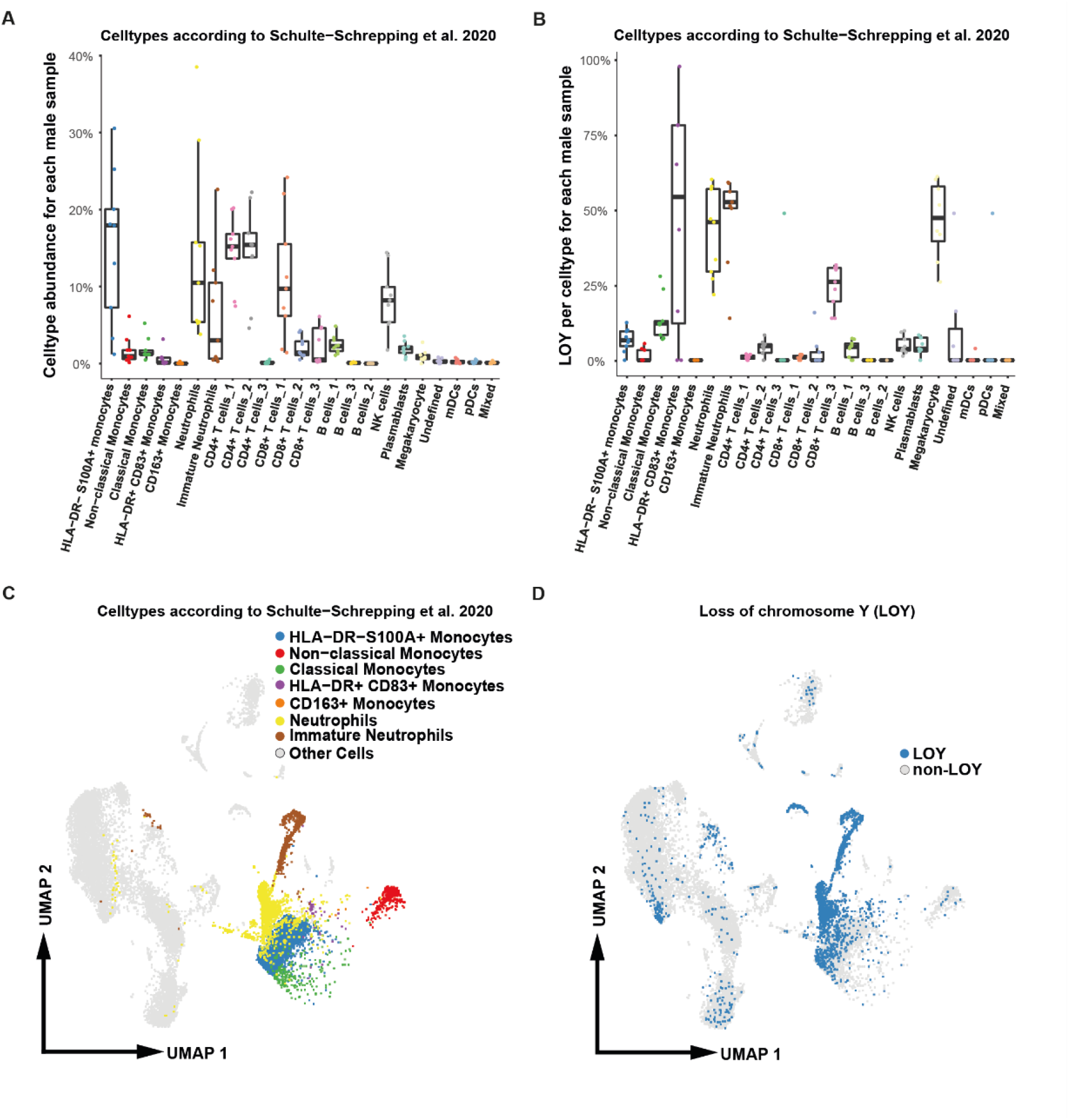
Distribution of cell types and cells with loss of chromosome Y (LOY) in PBMCs from critically ill patients with COVID-19. The dataset comes from Schulte-Schrepping et al. 2020 (PMID: 32810438) and we used cell type annotation reported therein. Nine PBMCs samples from six male patients with WHO grade >= 5 were used for calculations. **A**) Proportion of cell types in PBMCs; each data point represents a single sample. **B**) Proportion of cells with LOY per cell type and per sample; each data point represents a single sample. **C**) UMAP projection of sc-RNA-seq profiles, selected cell types are colored. **D**) UMAP visualization of sc-RNA-seq profiles colored according to LOY-status. Cells classified as LOY-cells had no detectable expression from chromosome Y [5].

In order to extend the above initial analysis, we collected ∼16 ml blood from 139 critically ill male COVID-19 patients, treated at two ICUs at University Hospitals (Uppsala and Gothenburg) in Sweden. The criterion for inclusion in our study as a critical COVID-19 patient was treatment at ICU (WHO grade 6 or higher). The median age for this cohort was 65 years, ranging 19-86 years. We also collected history of smoking habits, comorbidity data, as well as numerous clinical and biochemical parameters recorded during ICU-treatment (**Table S1**). We further designed a new protocol for cell-sorting using FACS, allowing analysis of LOY in LDNs, granulocytes, monocytes and PBMCs as well as in whole blood for all patients. We also analyzed 38 healthy controls, which were used for cell-sorting, followed by LOY analysis (n=17), as well for single-cell transcriptome analysis (n=34). The median age of controls was 71 years, ranging 61-91 years. Moreover, we studied four male subjects with a milder course of COVID-19, i.e. hospitalized patients with confirmed SARS-CoV-2 infection but with no signs of severe disease at the time of sampling.

The number of LDNs varied most among all cell types that were analyzed. Using FACS, we counted LDNs in all critically ill COVID-19 patients. Their median number (per 10,000 FACS events, unadjusted for age and smoking) was 60, ranging 1-458. The corresponding range of numbers for LDNs in milder COVID-19 patients (n = 4) and healthy controls (n=17) were 1-73 and 4-70 per 10,000 FACS events, respectively. **Figure 2** shows medians, 80% and 95% highest density intervals (HDI) for LDN cell counts, adjusted for age, age^2^ and smoking. The LDN cell counts in critically ill patients were significantly higher compared to healthy controls (median 86.8, 95%-HDI 61.8–119.1, p=1e-9) and to milder COVID-19 patients (median 63.1, 95%-HDI 2.6–107.2, p=.0089).

**Fig. 2.**
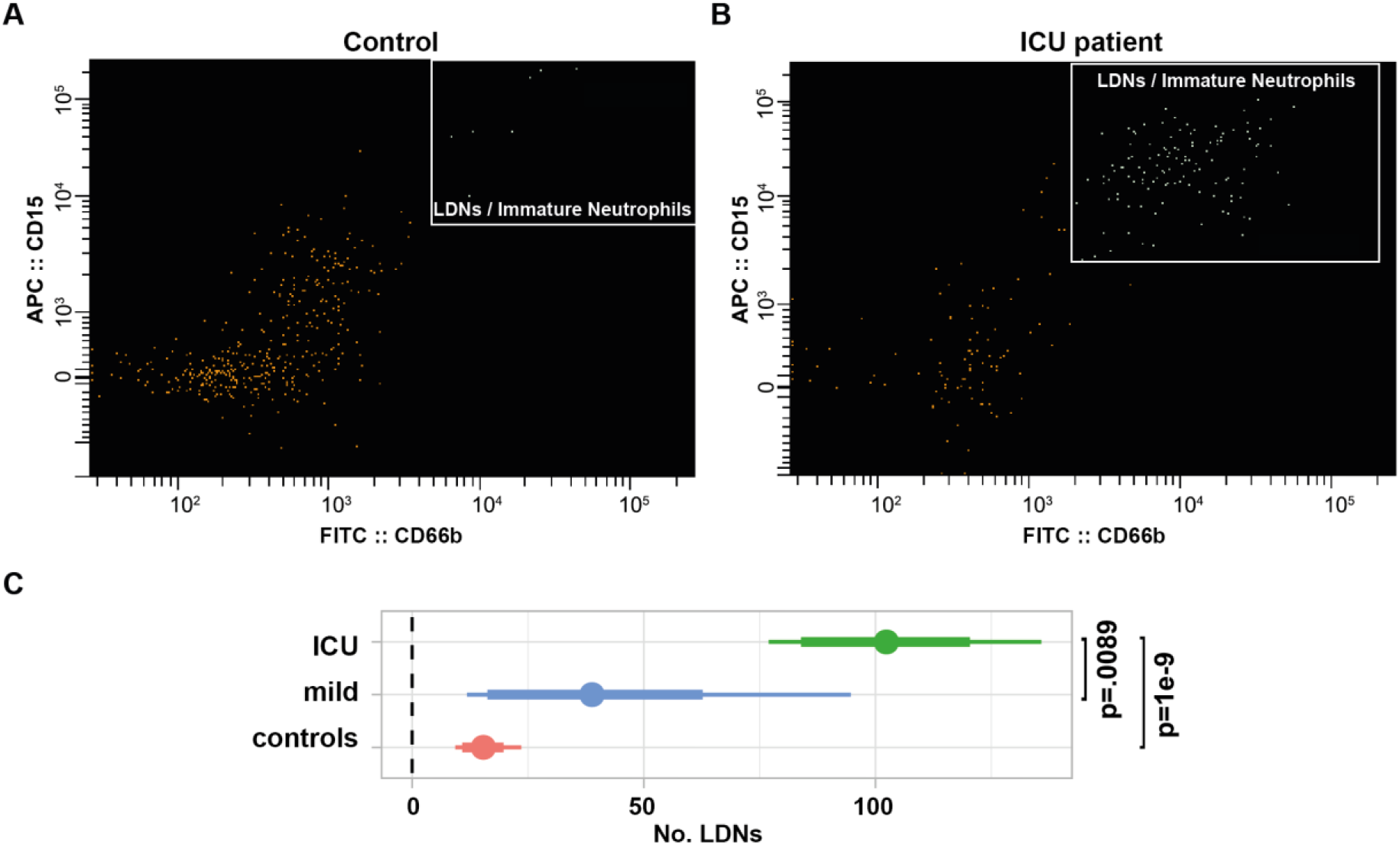
Differences in LDNs counts between controls and critically ill patients (treated at ICU). FACS images showing the gating strategy employed to identify low-density neutrophils (LDNs, also called immature neutrophils) for representative control (**A**) and ICU patient (**B**). PBMCs were gated for size and granularity first, LDNs were identified as CD15+ (APC) and CD66b+ (FITC) later. **C**) Comparison of the numbers of LDNs in PBMCs between critically ill patients (ICU, n=135), milder patients (n=4) and healthy controls (n=17). Points show medians; thick and thin horizontal bars show 80%- and 95%-HDI. P-values are shown after MVT correction.

The analysis of LOY status for all samples in this study was performed using droplet-digital PCR (ddPCR) as described [6]. The advantage of using ddPCR was a low input of DNA (∼50 ng) required to perform the LOY measurement, which was particularly important for analysis of LDNs from COVID-19 patients having low counts of these cells. Another advantage was a higher sensitivity in the lower range of measurements allowing to score LOY with 5% cut-off for cells with the mutation (**Fig. 3**). We were able to successfully detect LOY in DNA from LDNs sorted on FACS starting at about 10,000 cells. It should be stressed, however, that the number of successful measurements of LOY in LDNs was considerably lower than for other sources of DNA, due to the low number of LDNs from some COVID-19 patients. LOY percentages for all ICU-patients in five different cell populations (LDNs, granulocytes, monocytes, PBMCs and whole blood) were compared (**Fig. 3**). Adjusting for age, age^2^ and smoking the %LOY in LDNs was significantly higher than in monocytes (median 1.83%, 95%-HDI 0.94%–2.72%, MVT adjusted p=5e-4), in PBMCs (median 1.48%, 95%-HDI 0.58%–2.41%, p=0.013) and in blood (median 1.38%, 95%-HDI 0.5%–2.27%, p=0.018), and %LOY in granulocytes was significantly higher than in monocytes (median 0.75%, 95%-HDI 0.22%–1.26%, p=0.037). **Figures 3B** and **S2** show unadjusted values, also pointing to the highest level of LOY in LDNs compared with other cell types among critically ill patients. The range of cells with LOY also varied in different cell populations. The highest percentages of cells with LOY per subject in the five cell types were as follows: granulocytes, 85.5%; monocytes, 85.4%; whole blood, 81.7%; PBMCs, 51.5%; and LDNs, 36%.

**Fig. 3.**
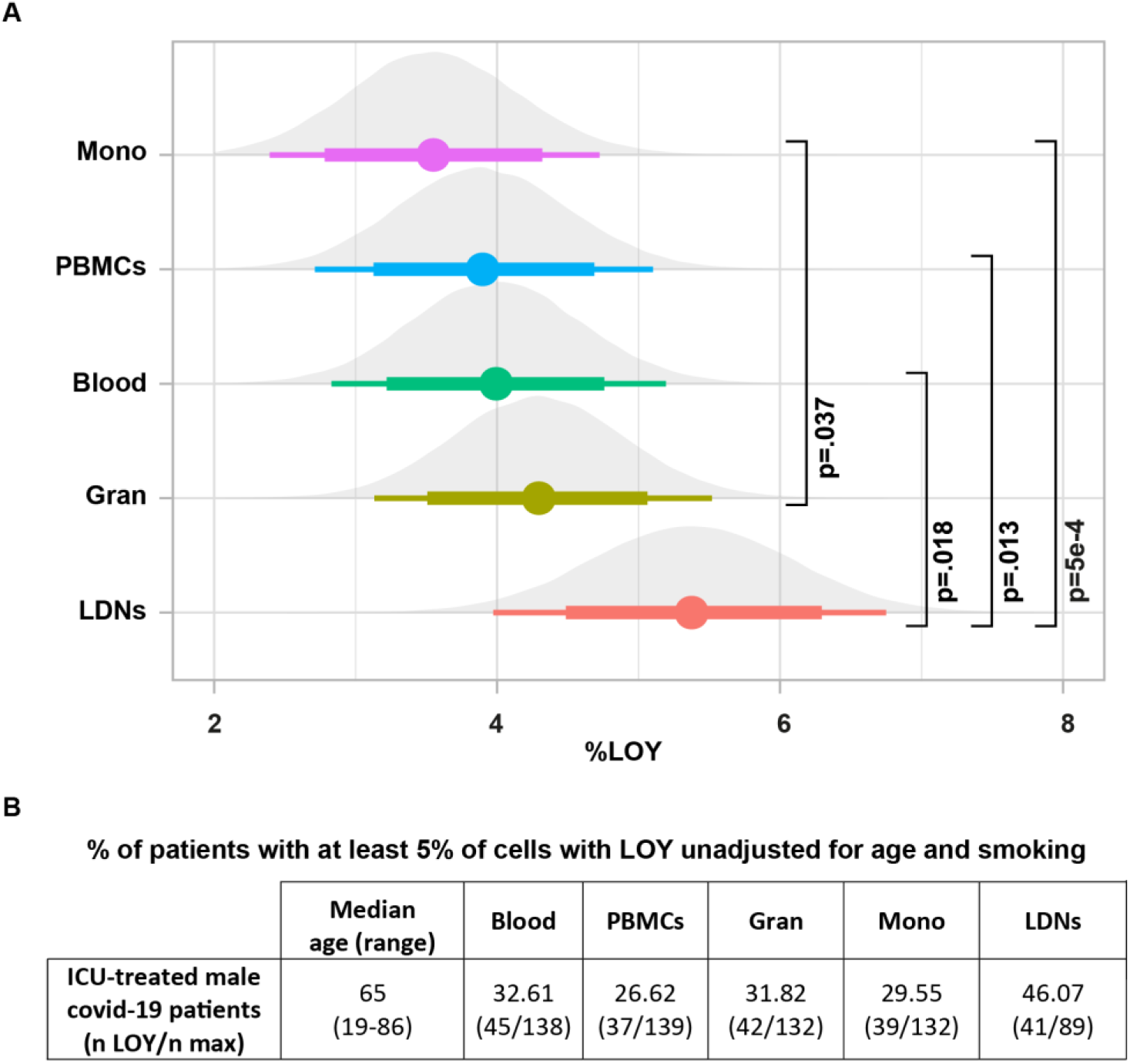
Comparison of %LOY across five cell populations for critically ill patients. **A**) Comparison of loss of chromosome Y percentages (%LOY) in monocytes, PBMCs, whole blood, granulocytes and low-density neutrophils (LDNs) within ICU-patients. Points show medians; thick and thin horizontal bars show 80%- and 95%-HDI. P-values are shown after MVT correction. **B**) Unadjusted percent of patients with LOY.

## LOY decreases in COVID-19 patient samples at the recovery stage

We further collected follow-up blood samples from 17 critically ill patients who recovered from the disease. The median number of days when the follow-up specimens were taken (from the date of first sampling) was 119 days, ranging 93-143. These samples were processed in the same way as described above for all critically ill patients. **Figure 4A** shows medians, 80%- and 95%-HDI for estimates of LDN cell counts, adjusted for age, age^2^ and smoking, demonstrating a radical decrease in the follow-up specimens (median 76.4, 95%-HDI 55.5-104, p=6e-11). Paired comparisons of cell numbers from LDNs and monocytes in 16 patients, where both ICU and recovery samples were available, also showed statistically significant decrease in the follow-up samples for LDNs (p=0.0066) and monocytes (p=0.013) using non-parametric paired Mann-Whitney-Wilcoxon test (**Fig. 4B**).

**Fig. 4.**
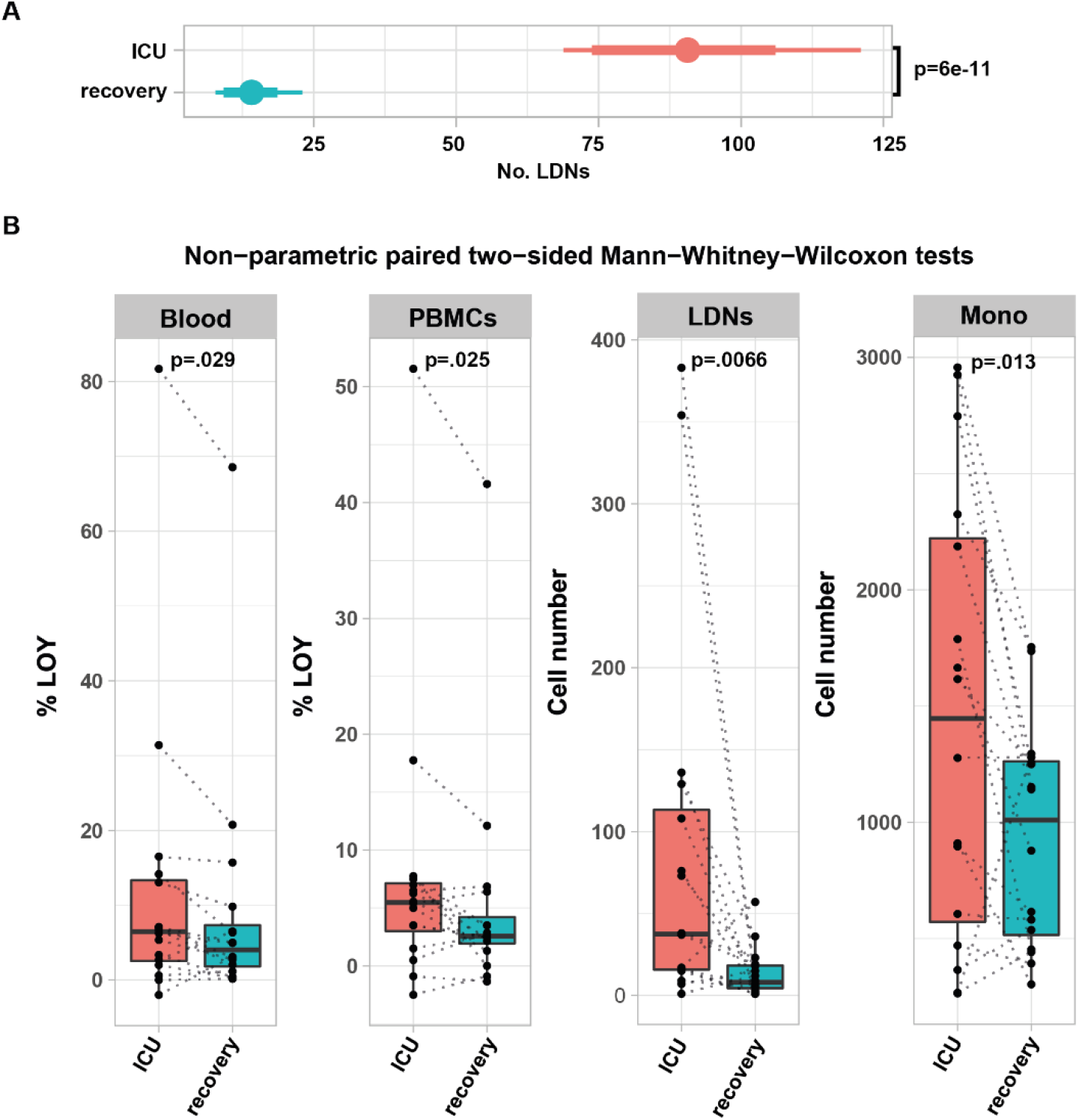
Leukocyte counts and LOY status for patients during ICU-treatment and during recovery. **A**) Comparison of the numbers of low-density neutrophils (LDNs) at intensive care unit (ICU, n=135) and 3-6 months after the discharge from ICU (recovery, n=17) in PBMCs. Points denote medians; thick and thin horizontal bars show 80%- and 95%-HDI, respectively. **B**) Paired comparison of loss of chromosome Y percentages (%LOY) in whole blood and PBMC, and numbers of LDNs and monocytes cells in PBMCs in 16 patients, where both ICU and recovery samples were available. Each dotted line connects the ICU and recovery data within the same patient. Boxplots show median, IQR as hinges and largest values no further than 1.5*IQR away as whiskers. Unadjusted p-values from paired two-sided Mann-Whitney-Wilcoxon non-parametric tests are shown.

We also performed paired LOY analysis in the five sorted populations of cells in 16 patients, where both ICU and recovery samples were available. Because the number of sorted LDNs from recovery samples were too low for DNA isolation (below 10,000 cells isolated via FACS from 16 ml of blood), we studied LOY only in the four other cell types; i.e. whole blood, PBMCs, granulocytes and monocytes. The %LOY values in DNA from whole blood (p=0.29) and PBMCs (p=0.25) decreased significantly in samples taken at the recovery stage using non-parametric paired Mann-Whitney-Wilcoxon test (**Fig. 4B**). In conclusion, the results from comparisons between ICU and recovery samples suggest the overall decrease of cell numbers that are mostly associated with the critical course of COVID-19 (LDNs and monocytes) as well as the overall decrease in the load of cells with LOY. The latter result underscores the dynamic character of aberrant cell clones with LOY that are present in COVID-19 patients.

## LOY correlate to clinical variables and co-morbidity in COVID-19 patients

Previous epidemiological studies showed an association of LOY with blood cell differentiation; that is thrombocyte- and erythrocyte cell counts were positively and negatively associated with LOY, respectively [29, 30]. In particular, binding sites of FLI1, a fate-determining factor promoting hematopoietic stem cell differentiation into thrombocytes rather than erythrocytes, showed a significant heritability enrichment of LOY GWAS signals using ChIP-seq data [29]. This effect of LOY might be important for the clinical outcome of critically ill male COVID-19 patients that display LOY, since increased counts of thrombocytes, hyper-coagulation and thromboembolic events are contributing to the high mortality in COVID-19 [21, 22]. We have therefore analyzed in our cohort of critically ill patients the correlation of LOY with the cell counts of thrombocytes and erythrocytes. For the patients treated at ICU in Uppsala, the cell counts for thrombocytes and erythrocytes as well as hemoglobin measurements were collected at the day of admission to ICU as well as the minimum- and the maximum values for these cells during the entire length of ICU stay. We estimated robust correlations of %LOY in five studied cell populations versus the above-mentioned clinical variables (**Fig. 5A** and **Table S1**). After adjusting for age, age^2^ and smoking status, the %LOY in PBMCs was significantly negatively correlated with maximum erythrocyte count (R = -0.25, 95%-HDI -0.45–-0.04) and with hemoglobin measurement, both at day 1 (R=-0.21, 95%-HDI-0.4–-0.007) and maximum values (−0.23, 95%-HDI -0.41–-0.02). In contrast, maximum thrombocyte counts were significantly positively correlated with %LOY in monocytes (R=0.023, 95%-HDI 0.02–0.42), granulocytes (R=0.023, 95%-HDI 0.02–0.43), and whole blood (R=0.24, 95%-HDI 0.04–0.43). Increased counts of thrombocytes in critically ill patients that have higher levels of LOY may predispose them for thromboembolic complications during ICU stay. Thus, we tested this variable against %LOY in blood and sorted subsets of leukocytes. Indeed, %LOY in LDNs (median 4.9%, 95%-HDI 0.29-9.6, p=0.041) and monocytes (median 2.7%, 95%-HDI 0.54-4.8, p=0.013) positively correlated with clinically confirmed thromboembolic events during ICU stay (**Fig. 5B**).

**Fig. 5.**
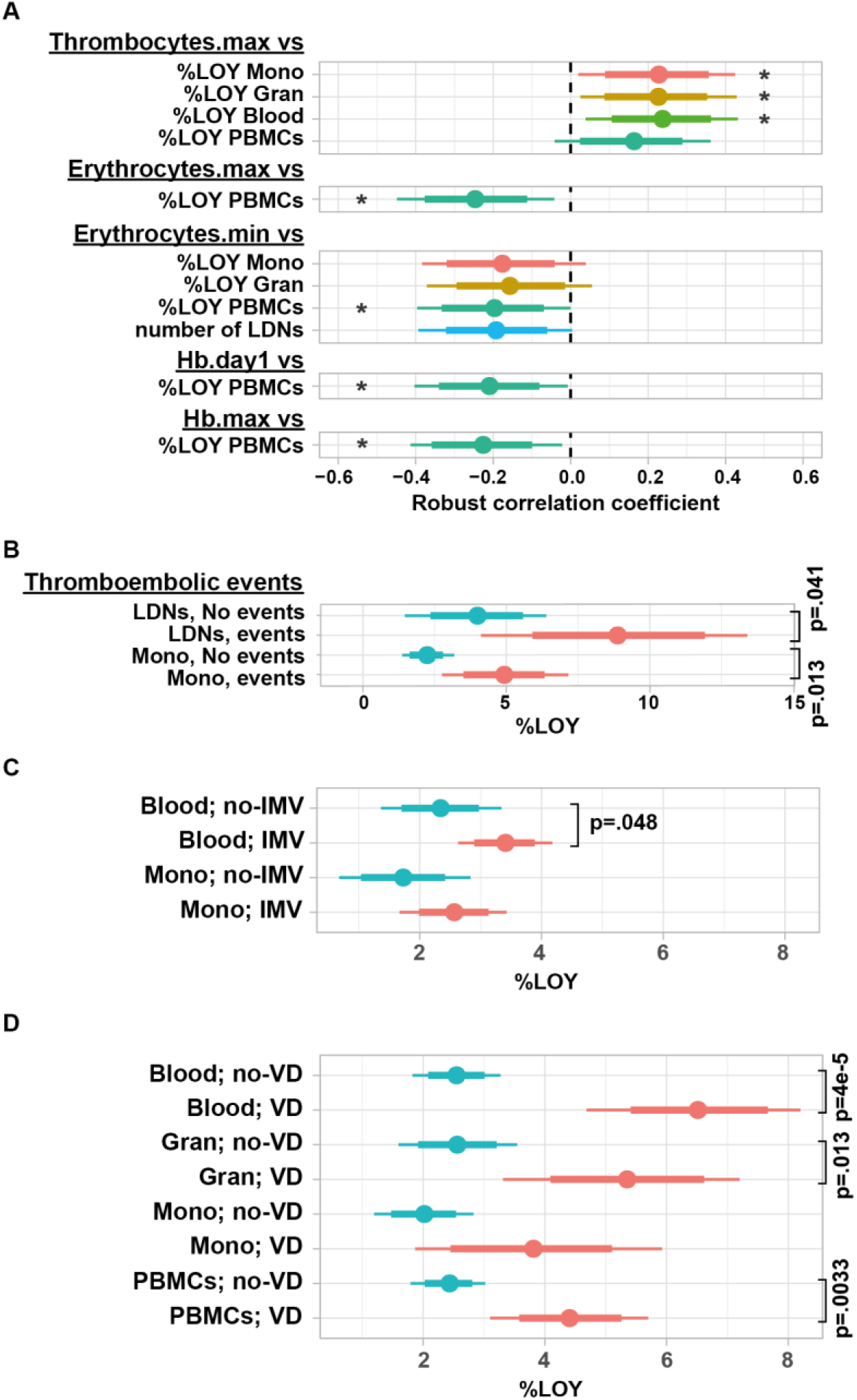
Clinical correlations. **A**) Correlation between %LOY vs thrombocyte, erythrocyte and hemoglobin (Hb) measurements. Robust correlation coefficients between %LOY in five types of cells and cell counts are shown; “.day1”, “.min” and “.max” denote levels at admission to ICU, as well as minimal and maximal measurements during ICU treatment, respectively. **B**) Comparison of %LOY in LDNs and monocytes with (n=20) and without (n=115) thromboembolic events at ICU. **C**) Comparison of %LOY in blood and monocytes at ICU, with (n=80) and without (n=56) invasive mechanical ventilation (IMV). **D**) Comparison of %LOY in four types of cells at ICU, with (n=26) and without (n=112) vessel disease (VD). Points show medians; thick and thin horizontal bars show 80%- and 95%-HDI, respectively. Asterisk denote that 95%-HDI is separated from zero and thus the correlation coefficient is statistically significant. Unadjusted p-values are shown.

The necessity to apply invasive mechanical ventilation (intubation and treatment in respirator) for some COVID-19 patients is one of the most severe consequences of this infection. We studied the possible association of this variable with %LOY, adjusting for age, age^2^ and smoking. We grouped our critically ill patients (**Table S1**) into those who did and did not receive this form of treatment. The %LOY in blood was significantly higher in patients treated with invasive mechanical ventilation (median 1.15%, 95%-HDI 0.04%– 2.3%, p=0.048) and showed a tendency in the same direction also in monocytes (median 0.99%, 80%-HDI 0.17%–1.77%) (**Fig. 5C**). This result suggests that %LOY might have a predictive value for the identification of patients that are at high risk for treatment with invasive mechanical ventilation and is another indication that LOY may contribute to severity of COVID-19.

We also tested a number of other clinical parameters (**Table S1**) that were collected for our critical COVID-19 cohort for correlation with %LOY, among them information about pre-existing comorbidities, prior to ICU admission. The co-morbidity, named “Vessel Disease (VD)”, was defined as the presence in the medical history of patients of any of the following conditions: coronary heart disease, stroke, diagnoses related to central vessels (aorta aneurysm, aorta stenosis, etc.) and diseases of peripheral vessels (intermittent claudication, skin ulcers of legs, etc). **Figure 5D** shows that, adjusted for age, age^2^ and smoking, the %LOY was significantly higher in VD patients in blood (median 4.0%, 95%-HDI 2.0%-5.8%, p=4e-5), in granulocytes (median 2.8%, 95%-HDI 0.52%-4.8%, p=0.013) and in PBMCs (median 2.0%, 95%-HDI 0.64%-3.26%, p=0.0033). Thus, we show that %LOY in blood is associated with common comorbidities present among COVID-19 patients treated at ICUs.

## sc-RNA-seq associates LOY and COVID-19 with a predominant down-regulation of genes in monocytes

We applied here single-cell transcriptomics using 10X Genomics platform for PBMCs from 30 males with critical COVID-19 and 34 healthy male controls to compare differentially expressed genes (DEGs) between cells with LOY and in normal state (abbreviated as non-LOY) in classical CD14+ monocytes. They were selected for the differential gene expression analysis because dysfunction of monocytes has been linked to pathogenesis of COVID-19 [18]. Furthermore, monocytes are abundant in the circulation of controls and expand in numbers in the COVID-19 patients. Yet another reason is our above described results showing that monocytes are frequently affected by LOY in COVID-19 patients and %LOY in monocytes displayed interesting correlations with clinical parameters. Cell types were annotated using Azimuth workflow as described [31, 32]. Cells were further classified as LOY-cells when they had no detectable expression from genes located in male specific region of chromosome Y (MSY) [5]. We performed three pairwise comparisons between gene expression levels at the baseline and three other groups. The baseline was the data from 22,676 non-LOY CD14+ monocytes from the 34 controls. This was compared with 3,741 LOY cells in the 34 controls (LOY effect), 41,486 non-LOY cells from 30 COVID-19 patients (COVID-19 effect) and with 6,015 LOY cells from COVID-19 patients (combined effect). We excluded MSY genes from these comparisons because their expression was absent in the single-cell data; i.e. this was the definition of LOY for single-cells [5]. The predominant pattern observed for top 500 DEGs was down-regulation of gene expression (**Fig. S3, Fig. S4** and **Fig. 6)**. The detailed results from the top 100 DEGs, including statistical significance for either down- or up-regulation in the three comparisons is illustrated in **Fig S4**. Similar analysis for top 500 DEGs is summarized in **Table S2**.

**Fig. 6.**
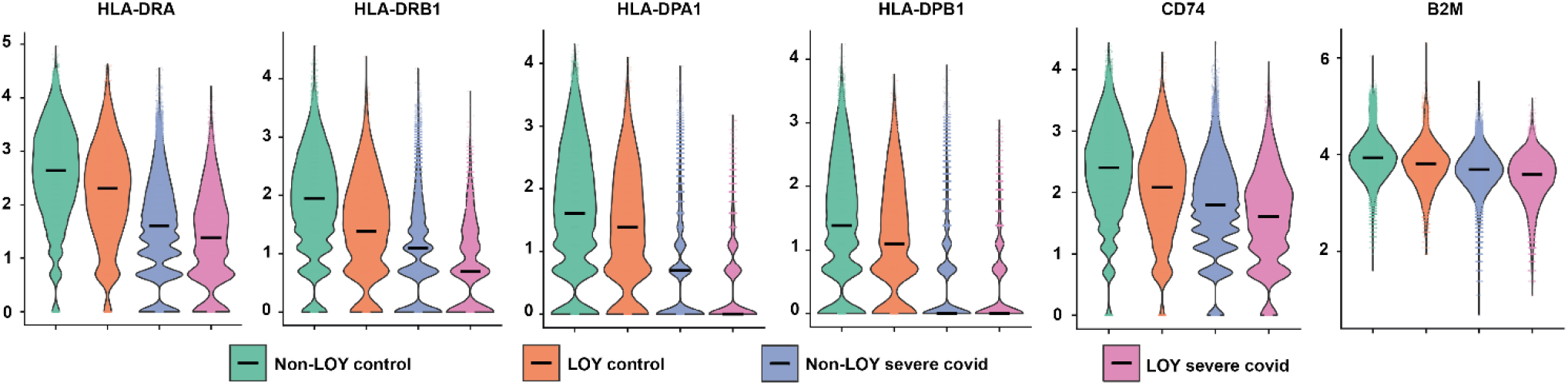
LOY- and covid-associated pattern of down-regulation of HLA-related gene expression in CD14+ monocytes. Violin-plots and median expression levels are shown after normalization by *sctransform* from Seurat: cells without LOY from controls (green; “baseline”; 22,676 cells), LOY-cells from controls (orange; 3,741 cells), cells without LOY from critical COVID-19 patients (blue; 41,486 cells) and LOY-cells from critical COVID-19 patients (magenta; 6,015 cells).

Furthermore, as is apparent from **Figs. 6** and **S4**, there was a dominant profile among down-regulated genes, with a consecutive and seemingly additive decrease of expression in each of the three pairwise comparisons. Genes showing a concordant down-regulation (average log fold-change <-0.1 in each of the three comparisons) were labeled as having a “down” pattern. There were 21 “down” pattern genes among the top 100 DEGs using this threshold. Similarly, genes showing a concordant up-regulation (average log fold-change >0.1 in each of the three comparisons) were labeled as having an “up” pattern. There were only six “up” pattern genes in the top 100 DEGs (**Fig. S4, Table S2**). The largest category of down-regulated genes was related to ribosome biogenesis/assembly, protein translation and protein targeting to membrane. Among the top 100 DEGs, there were 27 genes for ribosomal proteins and translation elongation or initiation factors. The second largest category of down-regulated genes among the top 100 (or top 500) DEGs are connected to antigen processing and presentation, predominantly via HLA class II genes and to a lesser degree through HLA class I system.

We further performed pathway enrichment analysis in Gene Ontology (biological process and molecular function) as well as in WikiPathway using 21 strictly defined “down” pattern genes from the top 100 DEGs (**Table S2**). As expected, top-enriched pathways were: translation (GO:0006412); peptide biosynthetic process (GO:0043043); MHC class II receptor activity (GO:0032395); CD4 receptor binding (GO:0042609); RNA binding (GO:0003723); MHC class II protein binding (GO:0042289); cytoplasmic ribosomal proteins (WP477); and pathogenesis of SARS-CoV-2 (WP4884). Essentially identical results were obtained using 58 strictly defined “down” pattern genes selected from the top 500 DEGs. Furthermore, there was no difference in results obtained when down-regulated MSY genes (due to LOY) were added into the enrichment analysis (**Table S2**). The corresponding analysis of up-regulated genes in Gene Ontology: Biological Processes (using six strictly defined “up” pattern genes from the top 100 DEGs, alternatively using 38 “up” pattern genes from the top 500 DEGs) revealed the following three most enriched pathways: neutrophil degranulation (GO:0043312); neutrophil activation (GO:0002283); neutrophil mediated immunity (GO:0002446) (**Table S2**).

## Discussion

The COVID-19 pandemic is the most serious new global disease of this century. Understanding the male bias in severity and mortality of COVID-19 is a central but poorly explored aspect of COVID-19 pathogenesis, which might be important for future strategies aiming at better prevention and treatment of critically ill patients [27]. We suggest here that LOY might be a factor underlying the higher severity and mortality of this infection in men. This would be another example of an age-related disorder associated with LOY, but the first acute, often life-threatening viral infection, representing a step forward in the understanding of the role of LOY in the susceptibility to disease, especially in relation to the dysfunction of the immune system. Male predominance for critical COVID-19 is consistent with a similar bias for prior SARS and Middle East respiratory syndrome epidemics (caused by SARS-CoV and MERS-CoV viruses, respectively) [33-35]. Moreover, male-female differences have also been reported for seasonal influenza A virus infections in Australia and Japan [36, 37]. Consequently, the importance of our results might not be limited to COVID-19 and this should be studied further in COVID-19 as well as other common viral infections.

We have recently reported [5], and show it here again in the context of COVID-19 that LOY can be scored on single-cell RNA-seq data, allowing analysis of the impact of this large mutation on transcription in selected leukocyte subpopulations. Using this approach, we re-analyzed published sc-RNA-seq data [18] showing that LDNs (or immature neutrophils), which play a key role in the pathogenesis of COVID-19, exhibit high levels of LOY in critically ill COVID-19 males (**Fig. 1**). It should be mentioned here that sc-RNA-seq has frequently been used in studies of COVID-19 and there exist numerous additional publicly available sc-RNA-seq datasets, which will facilitate follow-up and extension of our results in additional cohorts. Our results from sc-RNA-seq were confirmed at the DNA level, as we demonstrated that LDNs from critically ill patients displayed LOY, representing 46% of the studied COVID-19 patients (**Fig. 3**). Comparing the literature and considering the young age of patients in our cohort (median age 65 years), this is a very high number of subjects showing LOY in bulk DNA. This observation is also in line with other results suggesting that the myeloid lineage is the most affected by LOY [5, 38].

Epidemiological studies have associated LOY with myeloid cell differentiation [29, 30]. A high neutrophil to lymphocyte ratio is also a recognized marker of COVID-19 severity [39]. Considering the high percentage of LOY cells among neutrophils of COVID-19 patients, it is possible that clonal expansion and functional disturbance of these cells might be a consequence of LOY. We also demonstrated a decreased level of LOY in blood and PBMCs in recovering patients, suggesting that LOY could be a biomarker of severe disease.

Neutrophils are the cell type with the second highest daily turnover (after erythrocytes) and are intensively produced during infections [40]. Therefore, neutrophils are most prone to show clonal hematopoiesis of indeterminate potential (CHIP), which is associated with age-related accumulation of various post-zygotic mutations, among them LOY. In line with the above, we have recently shown that LOY in monocytes is accompanied by pathogenic somatic mutations in genes associated with DNA methylation, transcription regulation and DNA repair [38].

Our sc-RNA-seq results show that many members of HLA class II genes and *CD74*, as well as one crucial component of HLA class I system (beta-2 micro-globulin, *B2M*) are among the top down-regulated genes (**Figs. 6** and **S4, Table S2**) in CD14+ monocytes of COVID-19 patients. It is noteworthy that *CD74* was recently reported as factor that can block the entry of the SARS-CoV-2 and other viruses [41]. We have also recently shown that LOY is associated with dysregulation of autosomal genes and one example was *LAG3*, which is a component of HLA class II system [5]. Furthermore, previous work on immuno-phenotyping of leukocytes in critically ill COVID-19 patients showed down-regulation of some HLA class II proteins [42, 43]. Thus, our results together with published data suggest that LOY can negatively affect the immune system and could be responsible for dysregulation of numerous immune genes [5, 15].

We report correlations of LOY to clinical variables in critically ill patients, such as increased thrombocyte counts, which is in line with previous reports from non-covid cohorts [29, 30]. Thromboembolic complications also correlated with increased levels of LOY in LDNs and monocytes, which may contribute to the severity of COVID-19 disease in males with LOY in these cell fractions, eventually leading to organ failure, in particular affecting lungs. A recent paper [44] shows that thrombocytes from COVID-19 patients secrete increased levels of S100A8/A9 proteins forming heterodimer protein calprotectin, inducing the expression of coagulation-associated genes in vascular endothelial cells. The *S100A8* gene is the top upregulated gene in monocytes with LOY from COVID-19 patients (**Fig. S4** and **Table S2**) and this might represent another link between LOY and increased severity of COVID-19. In conclusion, LOY might have a predictive value for identification of patients that are at high risk for severe course of COVID-19 and this should be studied in larger cohorts. While the detailed mechanism of how LOY is involved in clonal expansion and disturbed function of myeloid cells remains largely unknown, our results support a link between LOY and emergency myelopoiesis and the role of LOY in modulation of COVID-19 disease severity, which ought to stimulate further research.

## Data Availability

All data produced in the present study are available upon reasonable request to the authors.

## ACKNOWLEDGEMENTS

We thank all the anonymous COVID-19 patients and volunteer healthy controls for acceptance to participate in the study, sample contribution and information provided in the medical questionnaire. We thank Maria Lindström, Anna Beskow, Susanne Aldén and the lab team at the Uppsala Biobank for their support with preparation of COVID-19 samples from patients. Uppsala Clinical Research Center (UCR), part of Uppsala University and Uppsala University Hospital, are also acknowledged in this respect. We thank the Uppsala Intensive Care COVID-19 research group; Tomas Luther, Anders Larsson, Katja Hanslin, Anna Gradin, Sara Galien, Jacob Rosén, Sara Bülow Anderberg, Labolina Spång, Erik Danielsson, Philip Karlsson and Amanda Svensson for the help with patient recruitment and sample collection. We acknowledge the Biobank West, a part of Sahlgrenska University Hospital and Gothenburg University for support with preparation of COVID-19 samples from patients recruited in Gothenburg. We thank Dr. Jakub Mieczkowski for bioinformatic advice. This study was supported by grants from the Science for Life Laboratory/KAW via National COVID-19 research, Swedish Heart-Lung Foundation, Swedish Cancer Society, the Swedish Research Council, Hjärnfonden, Alzheimerfonden, Konung Gustav V:s och Drottning Viktorias Frimurarestiftelse, and the Foundation for Polish Science under the International Research Agendas Programme (grant number MAB/2018 /6; co-financed by the European Union under the European Regional Development Fund) to J.P.D.; SciLifeLab/KAW national COVID-19 research program project grant to MH (KAW 2020.0182 and KAW 2020.0241), the Swedish Heart Lung Foundation to M.H. (20190639, 20190637 and 20210089) and the Swedish Research Council [2014-02569, 2014-07606, and 2016-02606 (the latter to J.D.J.)] as well as The Swedish Kidney Foundation (F2020-0054) to R.F. This study was also supported by grants from the SciLifeLab National COVID-19 Research Program, financed by the Knut and Alice Wallenberg Foundation [2020.0182 and 2020.0241, the Swedish Research Council (#2021-06545)] and the Swedish State Support for Clinical Research (ALFGBG-717531) to M.G. J.O.W was supported by KAW Foundation as part of the National Bioinformatics Infrastructure Sweden at SciLifeLab. Single cell RNA sequencing was performed at SNP&SEQ facilities at Uppsala University and SciLifeLab. SNP&SEQ is part of the National Genomics Infrastructure (NGI) Sweden supported by the Swedish Research Council and the Knut and Alice Wallenberg Foundation. Support by NBIS (National Bioinformatics Infrastructure Sweden) is gratefully acknowledged. Data handling and computations were enabled by resources provided by the Swedish National Infrastructure for Computing (SNIC) at Uppsala Multidisciplinary Center for Advanced Computational Science (UPPMAX) partially funded by the Swedish Research Council (#2018-05973).

## Materials and Methods

### Study design, sample collection and clinical variables

Male patients with COVID-19 were enrolled in Sweden, at Uppsala University Hospital (as part of the PronMed-study) and Sahlgrenska University Hospital in Gothenburg with confirmed positive SARS-CoV-2 nasopharyngeal test verified by RT-PCR. The COVID-19 cohort included 139 critically ill patients admitted to ICU, 4 milder patients (not requiring corticosteroid treatment, ICU-care, or high-flow oxygen at time of sampling) and 17 after recovery 3-6 months after discharge. Samples from COVID-19 patients were collected between July 2020 and August 2021 in Uppsala and Gothenburg, Sweden. Control samples were collected from 38 healthy individuals, in Gdansk, Poland. Informed consent was obtained from all the participants, or from next of kin for ICU-admitted patients, who were not able to give consent themselves. The research was approved by the local research ethics committee in Uppsala and Gothenburg, Sweden Dnr. 2017/043 with amendments 2019/00169, 2020-01623, 2020-20719 and 2021-01469; Dnr. 2020-01771 (COVID-19 cohorts) and Dnr. 2021-02205 (COVID-19 recovery samples). The Bioethics Committee at Medical University of Gdansk Dnr. NKBBN/564/2018 approved sample collection for control cohort in Poland. The study was conducted according to the guidelines of the Declaration of Helsinki. The full set of clinical variables that was used for statistical tests is shown in **Table S1**.

### Sample preparation

16 ml of blood was collected into BD Vacutainer® CPT™ Mononuclear Cell Preparation Tubes (BD). 1 ml of whole blood was saved for further DNA analysis. The tube was spun down within 4 hours of collection; PBMC fraction was washed with PBS and cryopreserved in FBS (Fetal Bovine Serum) for long time storage at -150°C. Granulocytes together with red blood cells were recovered from the fraction below the gel of the CPT tube. The red blood cells were then lysed twice with 40 ml of 1x RBC lysis buffer (PharmLyse, BD) for 15 min at room temperature. The granulocytes were washed with PBS and cryopreserved in FBS (Fetal Bovine Serum) for long time storage in -150°C. Cryopreserved PBMCs were thawed and washed with PBS according to the 10X Genomics protocol for the handling of frozen cells. Cell number and viability was determined with Trypan blue and Countess II FL automated cell counter (Thermo Fisher). Approximately 200 000 cells were saved for DNA extraction.

### Preparation of cells for FACS

For FACS analysis, the PBMCs were mixed with FITC labeled CD66b clone G10F5 (BD), APC labeled CD15 clone HI98 (BD), PE-CF594 labeled CD14 clone MφP9 (BD) and incubated for 20 min at 4°C. After incubation, PBMCs were washed with PBS and cell pellets were resuspended in 1 ml of PBS containing 3 mM EDTA. Before sorting, cells were filtered through 20 µm CellTrics strainer to remove aggregates. The target cell populations were isolated using FACS Aria III (Becton Dickinson). Data were acquired and analyzed using BD FACSDiva™ Software (Becton Dickinson). Live cells were sorted based on their FSC and SSC, a singlets gate based on FSC-H and FSC-A was used to remove doublet cells. Neutrophils were identified as CD15+ and CD66b+; monocytes were defined based on their size and as CD14 +. Cells were sorted to achieve a purity of at least 96%. After sorting, cells were centrifuged and cell pellets were stored in - 80°C freezer.

### DNA extraction

For samples with >50 000 cells the DNA was extracted using an in-house protocol previously described [5]. Cells were pelleted by centrifugation at 4000 rpm for 10 minutes and lysed with buffer containing 10 mM EDTA, 10 mM Tris–HCL (pH 7.9), 50 mM NaCl, and 1% N-Lauroylsarcosine sodium salt (Sigma) with 10 mg/ml proteinase K (Sigma). The DNA was then precipitated and resuspended in Low-TE. Samples with cell number between 10 000 to 50 000 cells were handled with an optimized cell lysis protocol. Lysis buffer without detergent was added to the cell pellet, together with 10 mg/ml proteinase K (Sigma) and incubated for 2 h in 50 °C. The cell lysate was then incubated in 95°C for 10 min to inactivate proteinase K. DNA from whole blood and granulocyte samples were extracted using QIAmp DNA Blood Midi kit (Qiagen).

### Analysis of the level of Loss of chromosome Y (LOY) using ddPCR

The LOY status was determined using ddPCR as described previously [6]. The DNA samples were pre-digested with HindIII (Thermo Fischer). Subsequently 50 ng of the digested DNA was used in the analysis. The digested DNA was mixed with PCR supermix for probes without dUTP (BioRad) together with primers and probes for the AMELX/AMELY TaqMan-assay, number C_990000001_10 (Thermo Fisher).

Quantification of the relative number of chromosomes X and Y in the sample was obtained by targeting the 6 bp sequence difference present between the AMELX and AMELY genes. Droplets were generated using the automated droplet generator (Bio-Rad), PCR was conducted using the T100 thermal cycler (Bio-Rad) and QX200 Droplet reader was used for the fluorescent measurements of droplets. The data was analyzed using the QuantaSoft software (Bio-Rad).

### Preparation of PBMCs for single cell RNA sequencing (sc-RNA-seq)

For sc-RNA-seq, samples from 30 individuals with critical COVID-19 and 34 controls were collected as described above. PBMCs were dissolved in a buffer containing PBS with 0.04% BSA at a concentration of 10^6^ cells/ml. The PBMCs were loaded on Chromium Next GEM Single Cell 3’ Reagent kit v3.1 (10x Genomics) for sequence library preparations according to manufacturer’s instructions. The single cell libraries were then sequenced using the NovaSeq 6000 and v1.5 sequencing chemistry (Illumina Inc.). The single cell library preparation and sequencing were performed at the Science for Life technology platform SNP&SEQ, Uppsala University, Sweden. CellRanger v6.0.1 (10x Genomics) was used to process sc-RNA-seq. To generate a digital gene expression (DGE) matrix for each sample, we mapped their reads to GRCh38 reference genome and recorded the number of UMIs for each gene in each cell. We excluded one control sample having more than 10 times the average number of reads and one critical covid sample having less than 40% fraction reads in cells.

### sc-RNA-seq data analysis of 10x Chromium data

sc-RNA-seq UMI count matrices were imported to R 4.1.0 and gene expression data analysis was performed using the R/Seurat package 4.0.4 [32]. We excluded cells based on the following quality criteria: more than 25% mitochondrial reads, less than 50 expressed genes or more than 7,500 expressed genes. We further excluded genes that were expressed in less than five cells. In addition, mitochondrial genes and MALAT1 have been excluded from further analysis. We removed doublets following R/DoubletFinder 2.0.3 best practice workflow [45].

We normalized count data using SCTransform function in Seurat, integrated data by embedding it into PBMC reference and annotated cell types using Azimuth workflow [32]. For two-dimensional data visualization we performed UMAP based on the first 50 dimensions of the supervised PCA data reduction. Cells classified as LOY-cells had no detectable expression from chromosome Y, according to Dumanski et al. 2021 [5].

### Differential expression tests

Differential expression (DE) tests were performed in CD14+ monocytes using FindMarker functions in Seurat with a hurdle model tailored to sc-RNA-seq data in MAST. Three pairwise comparisons were performed: group of cells without LOY from controls (baseline) vs. each of LOY-cells from controls (LOY), cells without LOY from critical COVID-19 patients (COV) and LOY-cells from critical covid patients (BOTH) groups.

Genes with >0.1 log-fold changes, expressed in >=20% of cells in >=1 tested groups, and Bonferroni-corrected p-values <0.05 in at least one pairwise comparison were regarded as significantly differentially expressed genes (DEGs). Top 500 DEGs ordered by adjusted p-values (breaking ties by log-fold changes) were summarized in **Table S2**.

### GO enrichment analysis

Genes showing concordant down-regulation (average log FC < 0 for all three comparisons) were named as showing “*down*” pattern. Among the top 100 significant DEGs, 21 DEGs showing a *down* pattern were taken as a gene set for enrichment test by R/enrichR package vs. GO Biological Process 2021 (GO:BP2021) and vs. GO Molecular Function 2021 (GO:MF2021). Since LOY-cells by definition have no gene expression from chromosome Y, only more conservative results involving genes not located on chromosome Y were reported. We noted that adding genes from chromosome Y into analysis or considering top 500 genes result in the same pathways showing significant enrichment, (see **Table S2** for details).

### Statistical analysis

Processing and statistical analysis was performed after completion of the experiments by the statisticians (D.S., J.O.W., A.S. and M.J.), who were not involved in execution of experiments. Therefore, the results of intermediate statistical analyses could not affect acquisition of experimental data. Data was analyzed using Bayesian regression models via full Bayesian framework by calling Stan 2.21 [46] from R 4.1 [47] using the brms 2.16 [48] interface. Predictors and outcomes were centered and scaled. To reduce the influence of outliers, Student’s t response distribution family with identity link function was used to model percent LOY outcomes and hurdle negative binomial distribution family with log link function was used to model FACS events counts. Models had no intercepts with indexing approach to predictors [49]. According to Stan recommendations [50] weakly informative priors were used for group-level effects, residual SD and group-level SD. P-values, adjusted using the multivariate t distribution with the same covariance structure as the estimates, were produced by frequentist summary in emmeans 1.6 [51]. Medians of the posterior distribution and 80%- and 95%-HDI were plotted. The contrast between groups was defined as significant if 95%-HDI did not include zero and adjusted P-value was ≤ 0.05. Moreover, FACS-sorting events count data and percent LOY data were analysed by Wilcoxon matched pairs signed-rank test. The significance level was set to p≤0.05.

## Data and Code Availability

Data supporting the findings of this study are available within the article, its Supplementary material or upon request. sc-RNA-seq data generated during this study are deposited at the European Genome-phenome Archive (EGA) under access number EGAS-nnn, which is hosted by the EBI and the CRG. Code is available on https://www.github.com/

